# Worsening of pre-existing psychiatric conditions during the COVID-19 pandemic

**DOI:** 10.1101/2020.05.28.20116178

**Authors:** Susanna Gobbi, Martyna Beata Płomecka, Zainab Ashraf, Piotr Radziński, Rachael Neckels, Samuel Lazzeri, Alisa Dedić, Asja Bakalović, Lejla Hrustić, Beata Skórko, Sarvin Es haghi, Kristina Almazidou, Luis Rodríguez-Pino, A. Beyza Alp, Hafsa Jabeen, Verena Waller, Dana Shibli, Mehdi A Behnam, Ahmed Hussain Arshad, Zofia Barańczuk-Turska, Zeeshan Haq, Salah U Qureshi, Ali Jawaid

**Author notes:** Equal contribution as first author (Susanna Gobbi, Martyna Beata Płomecka). Correspondence to: Dr. Ali Jawaid, MD, PhD.

## Abstract

This study anonymously examined 2,734 psychiatric patients worldwide for worsening of their pre-existing psychiatric condition during the COVID-19 pandemic. Valid responses mainly from 12 featured countries indicated self-reported worsening of psychiatric conditions in 2/3^rd^ of the patients assessed that was validated through their significantly higher scores on scales for general psychological disturbance, post-traumatic stress disorder, and depression. Female gender, feeling no control of the situation and reporting dissatisfaction with the response of the state during the COVID-19 pandemic, and reduced interaction with family and friends increased the worsening of pre-existing psychiatric conditions, whereas optimism, ability to share concerns with family and friends and using social media like usual were associated with less worsening. An independent clinical investigation from the USA confirmed worsening of psychiatric conditions during the COVID-19 pandemic based on identification of new symptoms that necessiated clinical interventions such as dose adjustment or starting new medications in more than half of the patients.

## Main Text

Corona virus disease 2019 (COVID-19) has emerged as the most critical global crisis of the 21st century. COVID-19 cases have exceeded 4.3 million as of May 15,2020.^1^ A number of studies indicate increase in symptoms of anxiety, depression, and other psychopathologies among the participants during the COVID-19 pandemic.^2–7^ In a previous study, we screened 13,332 individuals worldwide for general psychological disturbance, depression, and post-traumatic stress disorder (PTSD). Findings demonstrated increased odds for these conditions in individuals with pre-existing psychiatric conditions. Patients with previous history of psychiatric illness who reported worsening of their condition in response to the COVID-19 pandemic were identified as having higher risk for general psychological disturbance, contraction of the virus and relapse in condition.^8^ Worsening of psychiatric conditions is also associated with a higher risk of suicidal ideation, as demonstrated in the results of our study.^9,10^

Psychiatric conditions constitute a significant burden on health-care systems. Previous findings have associated these conditions with increased rates of mortality, disability and reduced overall economic output.^4,11–13^ Recently, the COVID-19 pandemic has given rise to even greater mental healthcare challenges for an already struggling system. Recent reports have called attention to the need for better mental health management of health-care workers, psychiatric patients, and general population.^3–6^ There has been an increase in utilization of mental health and suicide prevention helplines^14^ and new methods of psychological/psychiatric care delivery through telemedicine are being increasingly adopted.^15^ It is paramount for the optimization of mental health care delivery during these challenging times that the most vulnerable populations are identified and protected.

To address this, we performed a sub-analysis of data from participants with preexisting psychiatric conditions from our global study on the mental health impact of COVID-19. Each patient report of worsening of psychiatric conditions was then cross-analyzed with participants’ demographics, opinions/outlooks, personality traits, current household conditions, previous history, and other factors associated with COVID-19 to identify risk and resilience factors for worsening psychiatric condition. The results were then verified in an independent clinical cohort of psychiatric patients that consulted a psychiatry practice in Houston, TX, USA during the COVID-19 pandemic.

## METHODS

### Study Design

The study comprised two independent evaluations; 1. A cross-sectional electronic survey-based assessment of individuals above the age of 18 years willing to participate in the study, 2. Evaluation of anonymized clinical records of psychiatric patients above the age of 18 years.

### Online Survey

The anonymous online survey was conducted among participants from diverse demographic groups to examine the status of their pre-existing psychiatric conditions that were verified via standardized self-report scales for general psychological disturbance, risk for PTSD, symptoms of depression, and suicidal ideation. The survey was available online (placed on Google Forms platform) for a period of 15 consecutive days beginning 18:00 Central European Time on March 29, 2020 and concluding on 18:00 Central European Time on April 14, 2020.

The questionnaire was developed through close consultation between a neuroscientist, a neuropsychologist, a psychiatrist, a data scientist, and a psychiatry clinic manager. The questionnaire included closed-ended questions that assessed participant characteristics and opinions, and screened for neuropsychiatric conditions through standardized and validated self-report scales. The questionnaire prototype was prepared in English (Appendix 1) and translated into 10 additional languages (Arabic, Bosnian, French, German, Greek, Italian, Persian, Polish, Spanish, and Turkish; Appendix 2). The translation was performed by bilingual native speakers and vetted by volunteers native to those countries. The feasibility of each questionnaire was confirmed using pilot studies comprised of 10 participants each. These responses were excluded from the final analysis.

The questionnaires (Appendix 1) included a section on participant demographics (age, gender, country, residential setting, educational status, current employment status) household conditions (working/studying from home, home isolation conditions, pet ownership, level of social contact, social media usage, time spent exercising), COVID-19 related factors (knowing a co-worker, friend, or family member who tested positive for or demised due to COVID-19; prediction about pandemic resolution), personality traits (level of optimism, level of extroversion), previous history of psychiatric disease and/or trauma, previous exposure to human crisis, and level of satisfaction with actions of the state and employer during the current crisis. All questionnaires were rated on binary (yes/no) responses or Likert-type scales.

The other sections contained general health assessment based on WHO Self-Reporting Questionnaire-20 (SRQ), Impact of Event Scale (IES), and Beck’s Depression Inventory II (BDI).^16–18^ These scales were chosen based on their usage and efficacy in previously employed works studying the psychological impact of human crises such as the SARS epidemic.^19–27^ IES wording was purposefully adjusted to assess the impact of an ongoing event rather than a past event.

Using a non-randomized referral sampling (snowball sampling) method, participants were contacted by a team of 70 members (study authors and volunteers that have been acknowledged in the acknowledgement section) using electronic communication channels including posts on social media platforms, direct digital messaging, and personal and professional email lists. For the survey, 12 countries were included in the ‘featured’ list. These countries included USA, Spain, Italy, France, Germany, Iran, Turkey, Switzerland, Canada, Poland, Bosnia and Herzegovina and Pakistan. The data collection procedures were repeated at least thrice during the data collection period (March 29- April 14, 2020).

An overall total of 13,332 responses were collected. Surveys were excluded if they were completed by participants who were younger than 18 (n = 34), were missing responses for all dependent variables (n = 112), had been submitted previously (n = 325), were missing geographic location (n = 20), or were from WHO AFRO region (n = 24). When the responses were missing for individual items, the missing data were considered null and excluded from the analysis for that particular variable. In this follow-up study, however, only responses from participants who reported suffering from a previous psychiatric condition (n = 2,734) were considered valid.

### Clinical Study

The clinical data was extracted and analyzed for all the adult patients who consulted for online follow-up clinical evaluations at a Psychiatric care facility (Texas Behavioral Health) based in Houston, TX, USA during March 29- April 14, 2020. The inclusion criteria were previous diagnosis of major depressive disorder or anxiety disorders (generalized anxiety disorder and PTSD). Patients with diagnoses other than these and those under the age of 18 were excluded. Only the data from patients consenting to use of their records for this research were included in the study.

Clinical data for each patient was examined by clinic assistants blinded to the study design. The following information was extracted; age, gender, home-isolation status during COVID-19, social support during COVID-19, past exposure to trauma or a human crisis situation, and clinical diagnosis. Worsening of psychiatric conditions was assessed based on clinician report of new symptoms, need to increase or adjust the medication, and referral for a new therapy.

Data from 318 patients was considered valid for analysis. When the responses were missing for individual items, the missing data were considered null and excluded from the analysis for that particular variable.

### Ethical Considerations

Informed consent was obtained from each survey and clinical participant to allow anonymous recording, analysis, and publication of their answers. The data was collected in a completely anonymous fashion without recording any personal identifiers and the confidentiality of the participants was maintained throughout all phases of the study. The study procedures were reviewed and approved by University of Zurich Research Office for Scientific Integrity and Cantonal Ethics Commission for the canton of Zurich (Switzerland; Appendix 3), Nencki Institute of Experimental Biology, Warsaw (Poland; Appendix 4), and the Faculty of Medicine, University of Tuzla, Tuzla (Bosnia and Herzegovina; Appendix 5).

### Statistical Analysis

All statistical analyses were performed using R version v.3.6.3 and *Rstudio* (Rstudio team, 2015). All figures were produced using the packages *ggplot2* (Wickham et al., 2016) and *CGPfunctions* (Powell, 2020).

Unadjusted analysis for worsening of psychiatric conditions in both the survey and the clinical cohort involved Fisher’s exact test. For the survey, the categorical predictors included gender, residential status, education level, employment status, being a medical professional, working remotely from home, satisfaction with employer, satisfaction with the state (government), home-isolation status, interaction with family and friends, social media usage, ability to share concerns with a mental health professional, ability to share concerns with family and friends, prior exposure to a human crisis situation, previous exposure to trauma, level of extroversion, prediction about COVID-19 resolution and one’s self-determined role in the pandemic. For the clinical study, the categorical predictors included age, gender, home isolation status, and social support during home isolation.

Multiple logistic regression models were built to generate odds ratios (ORs) for worsening of psychiatric conditions both in the survey and the clinical cohorts. All statistical analyses were performed by the analysis team comprising MP, SG, PR, and AJ in consultation with ZB.

1. Chuck Powell (2020). CGPfunctions: Powell Miscellaneous Functions for Teaching and Learning Statistics. R package version 0.6.0. https://CRAN.R-
2. H. Wickham. ggplot2: Elegant Graphics for Data Analysis. Springer-Verlag New York, 2016.
3. RStudio Team (2015). RStudio: Integrated Development for R. RStudio, Inc., Boston, MA URL http://www.rstudio.com/.
4. Venables, W. N. & Ripley, B. D. (2002) Modern Applied Statistics with S. Fourth Edition. Springer, New York. ISBN 0-387-95457-0

## RESULTS

### Survey Study

#### Demographics

A total of 2,734 responses were considered valid with the highest responses from USA (874), Poland (255), Canada (246), Spain (205), and Pakistan (203). The distribution of the responses across the 12 featured countries and the WHO regions is presented in Supplementary Item S1. Canada had the highest (80.89%) proportion of patients reporting worsening of their psychiatric condition followed by Pakistan (72.41%), and the USA (67.5%). Turkey had the lowest percentage (28.57%) for worsening of psychiatric conditions (Main Item 1).

**Main Item 1.**
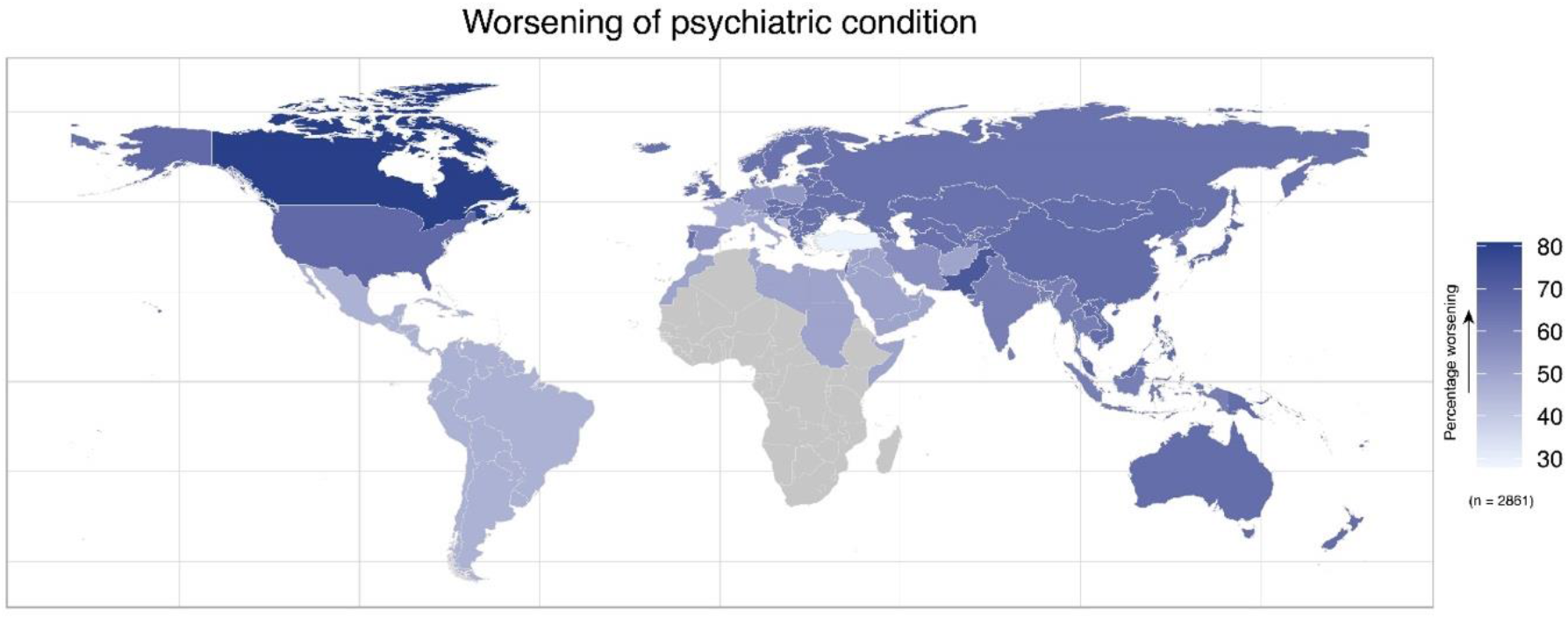
Geodemographic representation of the survey participants with pre-existing psychiatric condition that reported worsening of their condition. The map shows the percentage of worsening pre-existing psychiatric conditions separately for each of the featured countries, and for each of WHO regions.

There was a disproportion in valid responses, with higher numbers from those participants who were female (79.44%), residing in urban areas (84.6%), with advanced educational qualification, i.e., bachelor’s degree or higher (71.5%), working/studying remotely from home (65%), and currently under home-isolation with a partner/family (82.77%). Also notable were responses expressing some level of satisfaction with COVID-19-related employer (52.67%) and state response (64.26%) and spending less than 15 minutes on daily physical exercise (52.99%). A majority of participants also reported increased social media usage (65.42%), less-than-usual or minimal interaction with family and friends (64.88%), and feeling some level of control in protecting themselves and others during the COVID-19 pandemic (94.36%).

Participants’ report of worsening of psychiatric conditions was verified by comparing the SRQ, IES, and BDI and BDI scores between patients reporting worsening of psychiatric conditioning versus those reporting no change. All scores were significantly (p<0.05) higher in patients reporting worsening of psychiatric conditions. Distribution of patients reporting no change in their condition in comparison to worsening along the SRQ, IES, and BDI scaled further confirmed this pattern (Main Item 2).

**Main Item 2.**
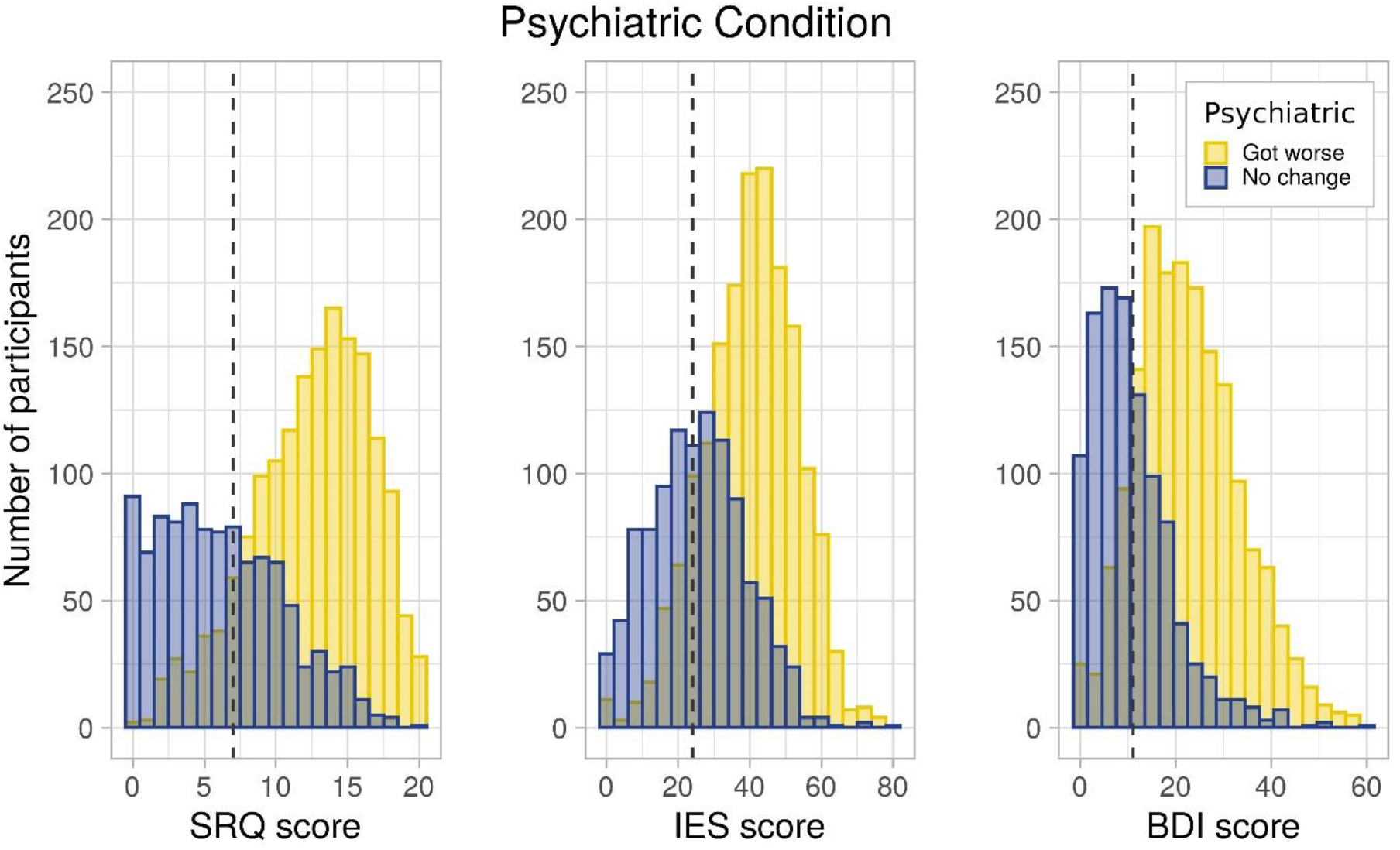
Population distribution of people with pre-existing psychiatric condition across SRQ, IES, and BDI score.

#### Unadjusted analysis of the worsening of psychiatric condition

Unadjusted Chi-square analysis of association between different patient factors and their report of psychiatric condition worsening revealed significantly higher reports of worsening in women, patients with advanced education, patients who reported being home isolated, and those with previous trauma exposure. Moreover, patients reporting dissatisfaction with the response of their government and employer during COVID-19 were more likely to report worsening of psychiatric condition. Finally, patients who identified themselves as a pessimist, felt lack of control during the current situation and had negative prediction about COVID-19 resolution were more likely to report worsening of their psychiatric condition.

On the contrary, patients that were able to interact and share concerns with their family and friends or with a health professional like usual during COVID-19 were less like to report worsening of their pre-existing psychiatric conditions. The details of the unadjusted categorical analysis are present in Main Item 3.

**Table 3.**
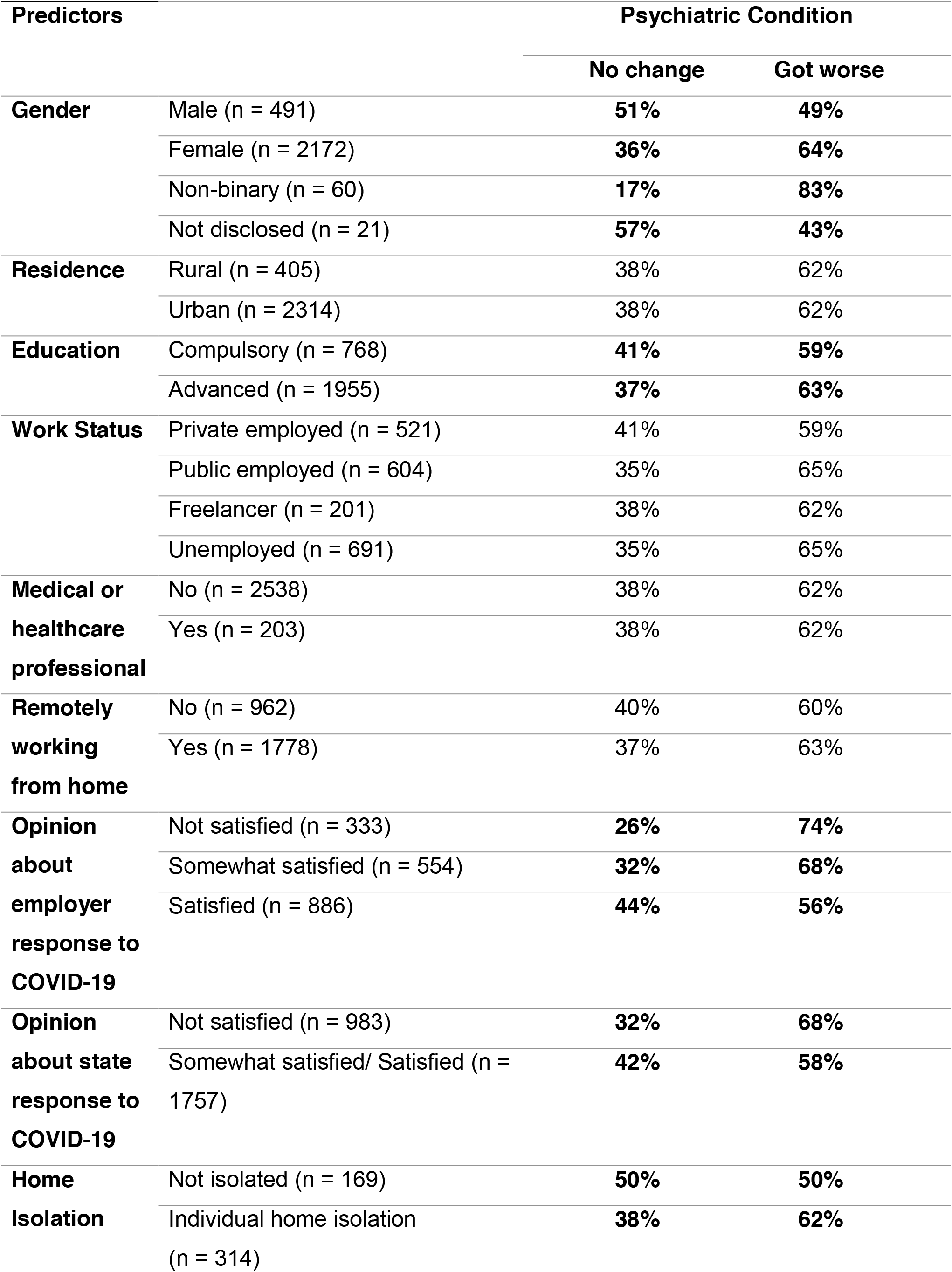
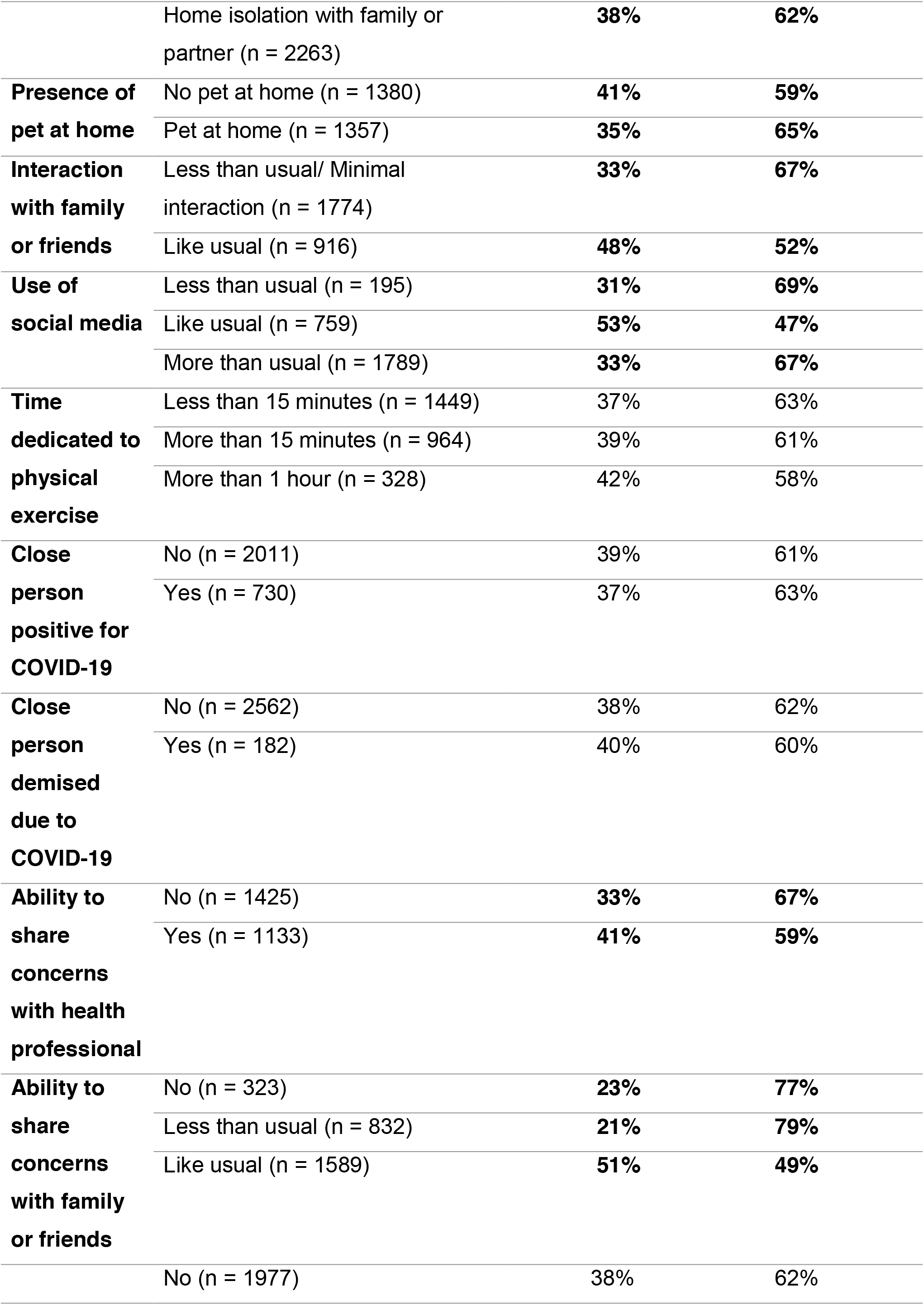
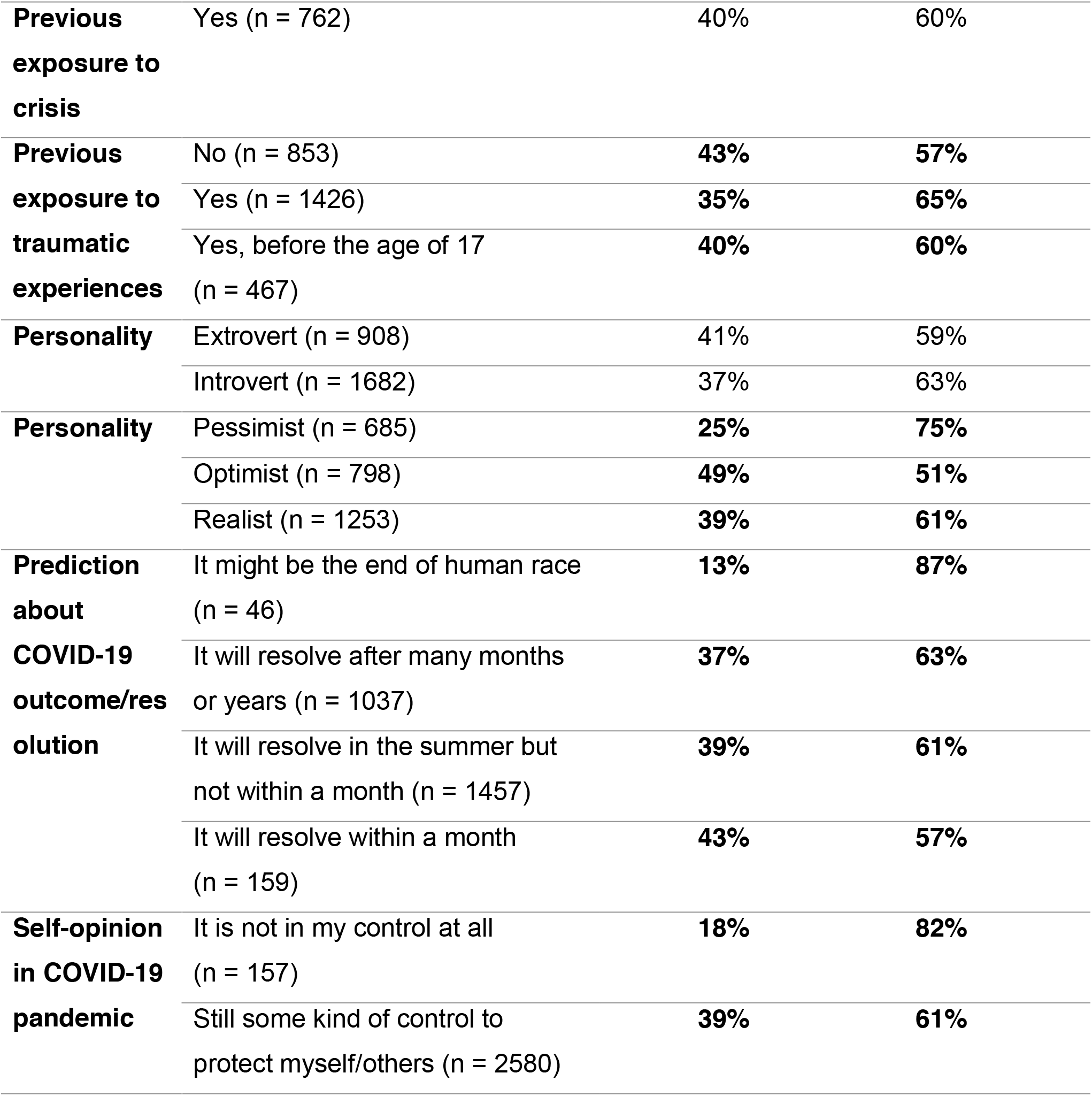
Association of psychiatric condition worsening and patient demographics/characteristics. This table shows the percentage of participants with and without a worsening or their psychiatric condition divided according to their demographics/ personal traits. The values are compared through an unadjusted Chi-squared test, and significant differences (p<0.05) are highlighted as bold. Specifically, each bold association indicates a difference in categories reported in the predictors’ column vertically.

#### Adjusted analysis of the worsening of the psychiatric condition

Adjusted analysis was then performed for patients’ report of psychiatric condition worsening via logistic regression to adjust for confounding associations. Report of feeling no control over the situation during the COVID-19 pandemic showed an 89% increase in the odds of reporting worsening of psychiatric condition (OR: 1.89, 95% CI: [1.18, 3.03]). Similarly, no or minimal social interaction during COVID-19 was associated with higher odds of reporting worsening of the psychiatric condition during COVID-19 (OR: 1.56, 95% CI: [1.30, 1.87]). Not being satisfied with the government’s response also showed an increased probability of worsening of psychiatric condition during COVID-19 (OR: 1.31, 95% CI: [1.09, 1.58). Finally, female psychiatric patients were more likely to report worsening of their psychiatric condition compared to male patients (OR: 1.70, 95% CI: [1.28, 2.00], Main Item 4).

**Main Item 4.**
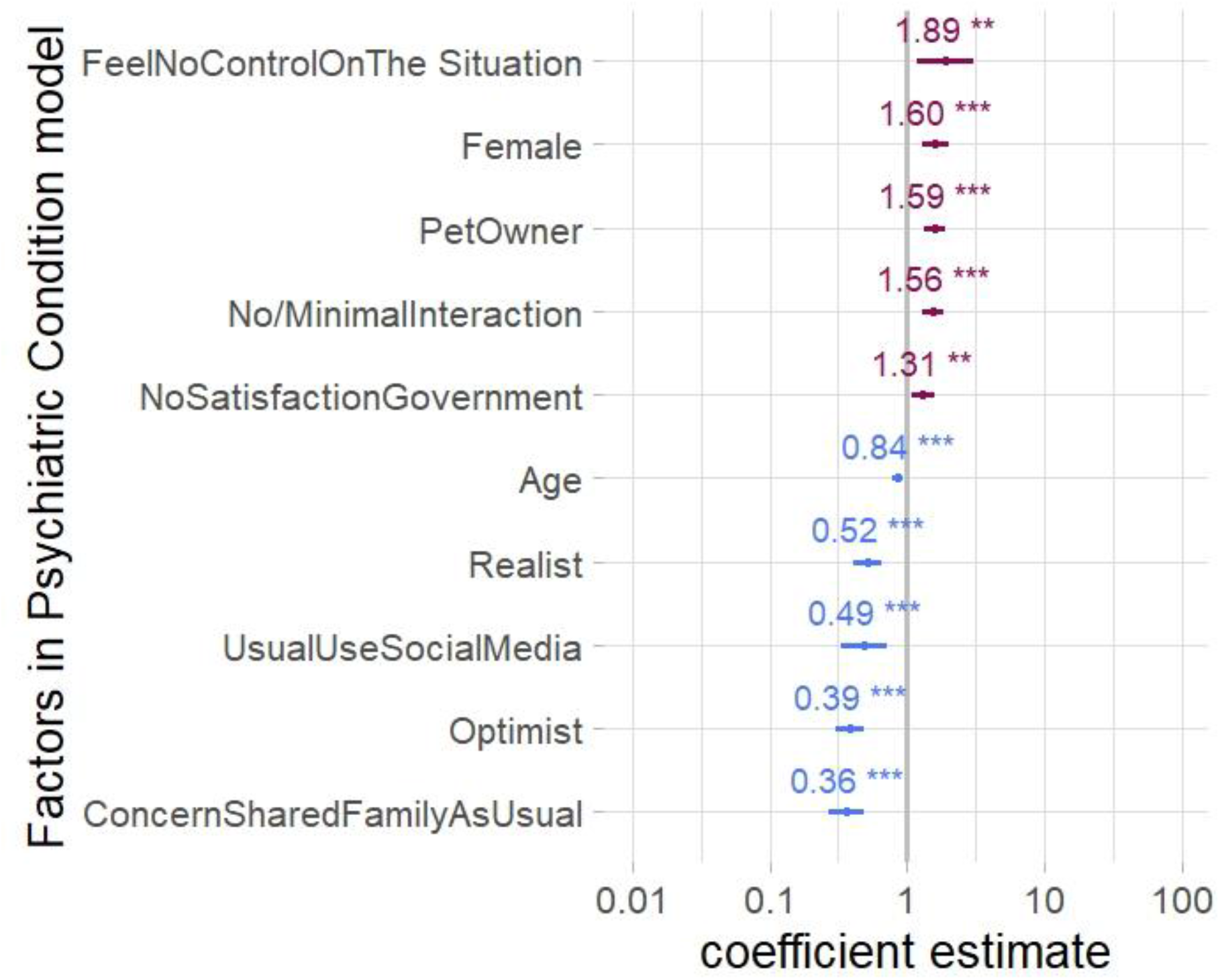
Factors associated with psychiatric condition worsening. Foster plot shows the mean estimates and the 95% confidence intervals (CI) for adjusted coefficients significantly affecting the reported worsening of psychiatric condition by the patients. Factors indicating more odds of worsening are shown in red while factors indicating less odds of worsening are in blue.

On the contrary, the ability to share concerns with family and friends like usual and optimistic attitude decreased the worsening of the psychiatric condition (OR: 0.39, 95% CI: [0.30, 0.49] and OR: 0.36, 95% CI: [0.27, 0.49]. Furthermore, as-usual usage of social media during COVID-19 and considering oneself a realist also decreased the probability of worsening of psychiatric condition (OR: 0.49, 95% CI: [0.34, 0.71] and 0.52, 95% CI: [0.41, 0.65].

#### Clinical Study

The valid clinical samples comprised 71.58% females and the diagnosis of a vast majority (83.56%) of patients was major depressive disorder. Clinicians identified new symptoms in 44% of patients, with sleep disturbance being the most common emerging symptom. Collectively, clinicians felt the need to make treatment adjustments in almost half of the patients in the form of starting a new medication or treatment modality or adjusting the dose of a currently prescribed medication. (Supplementary Item S3).

Among the patient-related factors, female gender significantly increased the likelihood of a change of medication by the clinician (OR: 2.22, 95% CI: [1.03, 4,49]). However, other patient-related factors such as age and level of social support during home-isolation wer e not associated with any clinical intervention (Main Item 5).

**Table 5.** Factor associated with clinician change of medication. Logistic regression analysis to assess the effect of patient gender, social support during home isolation, and age predicts increased likelihood of medication change by the clinician for female psychiatric patients during the COVID-19 pandemic.

|  | Change of<br>medication |
| --- | --- |
| Female | 2.22 * [1.03,4.79] |
| Social Support | 1.24 [0.57,2.70] |
| Age | 0.90 [0.66,1.21] |
| N | 291 |

\* p < 0.05.

### DISCUSSION

This study highlights a significant impact of the COVID-19 pandemic on psychiatric patients worldwide. At least 50% of the psychiatric patients evaluated in this study from 8 of the 12 featured countries reported worsening of psychiatric conditions. Notably, the self-reported worsening of psychiatric conditions was cross-validated with patients’ scores on validated scales assessing general psychological disturbance, risk for PTSD, and depression. Severity of psychopathology assessed through these scales confirmed the patients’ report of psychiatric condition worsening. Finally, clinician-reports from an independent cohort of psychiatric patients in the US confirmed that more than half of the patients reported new symptoms and required treatment adjustments during the COVID-19 pandemic.

In addition to ascertaining if there has been a general worsening of psychiatric conditions during COVID-19, a major aim of this study was to identify risk factors for such worsening. Female gender, having no or minimal interaction with family and friends, not being satisfied with the actions of the government, and feeling lack of control over the situation were associated with worsening of psychiatric conditions in the survey cohort. Patients who were older, considered themselves optimists or realists, used social media like usual, and were able share their concerns with family and friends during COVID-19 like usual were less likely to report worsening of psychiatric conditions. Notably, examination of the clinical cohort confirmed some of these findings. Clinicians reported significantly higher adjustment of medications for female psychiatric patients.

To the best of our knowledge, this is the first systematic assessment of psychiatric patients during the COVID-19 pandemic. The results of this study confirm previous speculations and concerns about the vulnerability of this population during this crisis.^7,28^ Compared with previous studies on the impact of mental health during pandemic, this study focuses on the deterioration of psychiatric illnesses in response to COVID-19. Other studies have focused on vulnerable populations including indigenous, migrant and imprisoned populations, people with disabilities, women,^29^ front-line workers^30^ and the elderly,^11^ but thus far has not included populations with pre-existing psychiatric illnesses. Tracing the worsening of psychiatric illnesses in response to COVID-19 can provide the insight necessary to improve mental health systems and the resources they can offer. Moreover, keeping the vulnerability of those with pre-existing psychiatric illness in mind, health systems can become better equipped to address the concerns of this population, mitigate the risk of further mental deterioration and reduce prevalence of suicidal ideation. Previous studies have reported the importance of adequate procedures to detect early mental health worsening,^31^ but have scarcely been conducted in the context of pandemics such as COVID-19.

Identifying factors that are associated with worsening of psychiatric conditions has important implications for psychiatric prognostics and therapeutics. In our previous study, patients with prior psychiatric disease reported increased suicidal ideation.^10^ Understanding factors associated with psychiatric disease during a pandemic can help the patients, their family, and care-givers to screen and identify those at an increased risk of mental health crises situations such as suicide attempts. Factors identified in this study including gender-based factors and prior exposure to trauma warrant further investigation to ensure that health systems can provide for the needs of a vulnerable population.

Previous research has highlighted increased gender-based disparity and violence associated with humanitarian crises.^32^ During the Ebola outbreak of 2014–16, women were increasingly at risk of abuse, violence and a lack of access to protective instructions.^29^ Moreover, existing gender norms and inequality can exacerbate the effects of economic insecurity, social-isolation, disaster-related unrest, reduced health service accessibility, inability to escape abusive partners, and violence against health-care workers for women. Measures such as social-isolation have increased women’s exposure to domestic violence: early reports from a police station in China’s Hubei Province recorded thrice the amount of domestic violence reports during the COVID-19 quarantine period of February 2020.^29^ Since women also have an increased risk of psychiatric disease such as depression and anxiety compared to men, the gender-based disparity and violence associated with the pandemic intersects with pre-existing conditions and puts women more at risk. Hence, governments and public health experts should recognize the needs of women and women with psychiatric diseases to counter the vulnerability and risk they face.

There are several strengths of our global and immediate approach to the examination of the vulnerable population of psychiatric patients during COVID-19. First, the sample size is large: participants from 12 countries responded to reliable measures to predict and analyze their mental well-being. Second, to avoid a weak external validity, the study was administered in 11 different languages, ensuring generalizability across countries and cultures. Participants from the 12 countries represented a diverse perspective according to the economic structure and government support provided by their respective countries. For instance, countries like Canada, France, Germany, Italy, Spain, Switzerland, and USA are classified as high-income economies according to the World Bank Atlas, whereas, Bosnia and Herzegovina, Iran, Pakistan, and Turkey are considered middle or lower-income countries.^33^ Third, as one of the earliest examinations of the mental health impact of COVID-19, our study carries the unique strength of immediate data collection during the peak of the COVID-19 pandemic in North America and Europe, between the dates of March 29 and April 14, 2020. Lastly, the results of our study isolate the worsening conditions of psychiatric disease—a novel contribution to the literature of pandemic research. The worsening of conditions assessed by clinicians in an independent cohort provides a precedent to address and prioritize mental health and is an important contribution and strength of this study. This study also has potential limitations that warrant consideration for the interpretation of results. First, the sampling method is non-randomized for the survey cohort. While a non-randomized approach has potential disadvantages, we hope that the results of this study can nonetheless serve as a resource and catalyst for further investigation. For a similar global or continent-wide study, entities such as the World Health Organization (WHO) and the EU (European Union) could develop and administer a similar study with a wider reach. Second, the data were exclusively collected online for the survey – this may have excluded those less well-versed in web-usage such as underdeveloped, rural or disadvantaged populations. Nevertheless, to counter existing language-barriers that may be furthered by computer illiteracy, we translated the survey in native and official languages for each of the featured countries. Lastly, a longitudinal assessment of the evolution of psychological symptoms in response to the COVID-19 pandemic is imperative and indeed, an on-going investigation by our group of researchers.

In conclusion, this effort highlights a significant impact of the COVID-19 pandemic on the mental health of psychiatric patients and elucidates prominent associations with their demographics, house-hold conditions, personality traits, and attitude towards COVID-19. These results could serve to inform mental health professionals and policymakers across the globe, aiding in dynamic optimization of mental health services during and following the COVID-19 pandemic, and reducing its long-term morbidity and mortality.

## Data Availability

All data presented in the main and supplementary
items are deposited on the repository below and are available for verification upon request.

https://osf.io/3vupe/?view_only=80f71b6f0c8d49b08573ea12eab10d33

## Acknowledgments

We gratefully acknowledge the contribution of Luciana Armengol (Argentina), Prof. Anthony Hannan, Maxine Mason, Qi Hui Poh (Australia), Taria Brkić (Bosnia and Herzegovina), Barbara Levinsky (Brazil), Alexandra Schimmel and Lea-Caya Bissonnette (Canada), Claudia Valenzuela Rios (Chile), Marc Scherlinger, Alice Tondre, Lola Kouroma, and Morgane Roth (France), Katharina Schlerka, Lisa Garrelts, and Romy Seifert (Germany), Lena Heck (Germany/ Switzerland), Varsha Hooda, Deepak Tanwar, and Chakradhar Yakkala (India), Prof. Mohammad Es haghi and Sepehr Namirad (Iran), Darren Kelly (Ireland), Nour Mosawy (Jordan), Dayra Lorenzo (Mexico), Chirine Katrib (Lebanon/ France), Usman Mukhtar, Uzair Jaswal, and Mubaris Bashir (Pakistan), Prof. Kornelia Kedziora-Kornatowska, Milena Czarnocka and Juli Davis (Poland), Ana Alexandra Moraru (Romania), Shoaib Jawaid (Saudi Arabia/ United Arab Emirates), Myriam Merarchi (Singapore/ France), Michelle McLuckie, Doman Obrist, Niharika Gaur and Graciela Huber (Switzerland), Aurelia Muller (Taiwan/ Germany/ Switzerland), Burak Ozan (Turkey), Carmen Neagoe and Aleena Malik (UK), Anastasiia Timmer (Netherlands/ USA), Colette Rausch, Prof. Paul Schulz, Prof. Mo Salman, Saleha Tahir, Laura Luebbert, Sarish Khan, Rebecca Sager, Lupita Lozano, and American Physician Scientist Association (USA) for their dedicated help in data collection. We are also thankful to Lena Heck and Giuseppe Parente (University of Zurich) for technical support. Finally, we would like to express our gratitude to Prof. Selmira Brkić (Faculty of Medicine, University of Tuzla, Bosnia and Herzegovina), Prof. Leszek Kaczmarek (Director, Nencki-EMBL Braincity, Warsaw, Poland), University of Zurich Research Office, Zurich Cantonal Ethics Commission, Texas Behavioral Health, and European MD-PhD Association for their expedited review of the study procedures under extra-ordinary circumstances and for their organizational support.

## Funding

The authors worked voluntarily for this project and have no funding source to disclose. AJ is supported by an International Research Agenda (MAB) grant by Foundation for Polish Science (FNP).

## Author contributions

MP and SG contributed in conceptualization, questionnaire development, data collection, data mining, data analysis, visualization, review and editing. RN contributed in data collection, manuscript writing, review and editing. BS, SL, KA, AD, AB, LH, SE, HJ, LRP, VW, BA, MB, and DS contributed in questionnaire translation, data collection, data mining, review, editing, and project co-ordination. PR contributed in data analysis and visualization. ZA contributed in data collection, manuscript writing, review, and editing. ZB contributed in data analysis. ZH, AHA and SUQ contributed in clinical data collection and project co-ordination. AJ contributed in conceptualization, questionnaire development, study approval, data collection, data analysis, data visualization, manuscript writing, review, editing, project administration and supervision. All authors have reviewed and approved the final draft.

## Competing interests

The authors declare no competing interests. request.

## Data and materials availability

All data presented in the main and supplementary items are deposited on the repository below and are available for verification upon request. https://osf.io/3vupe/?view_only=80f71b6f0c8d49b08573ea12eab10d33

## Notes

### Competing Interest Statement

The authors have declared no competing interest.

### Clinical Trial

This study doesn't include any clinical trial.

### Author Declarations

Informed consent was obtained from each survey and clinical participant to allow anonymous recording, analysis, and publication of their answers. The data was collected in a completely anonymous fashion without recording any personal identifiers and the confidentiality of the participants was maintained throughout all phases of the study. The study procedures were reviewed and approved by University of Zurich Research Office for Scientific Integrity and Cantonal Ethics Commission for the canton of Zurich (Switzerland), Nencki Institute of Experimental Biology, Warsaw (Poland), and the Faculty of Medicine, University of Tuzla, Tuzla (Bosnia and Herzegovina)

